# Evaluation of reproductive profiles, epigenetic aging, and mortality in post-menopausal women

**DOI:** 10.1101/2025.02.08.25321904

**Authors:** Qiaofeng Ye, Aaliya Ahamed, Idan Shalev, Laura Etzel

**Affiliations:** Department of Biobehavioral Health, The Pennsylvania State University, University Park, PA 16802, USA; Social Science Research Institute, Duke University, Durham, NC 27708, USA

**Keywords:** Hormones, Biology of Aging, DNA methylation

## Abstract

Evolutionary theories of aging indicate trade-offs between reproduction and longevity. Epigenetic clocks, such as PhenoAge, GrimAge, and DunedinPoAm, were designed to reflect biological age and be used as surrogates for mortality and healthspan. The current study investigated the connection between reproductive profiles, epigenetic aging and mortality among post-menopausal women (50-85 years) with data from the National Health and Nutrition Examination Survey across the United States (N=770). Using latent profile analysis, we identified four distinct reproductive profiles: high gravidity but average parity (Class 1); high gravidity and parity (Class 2); early menopause (Class 3); an average profile (Class 4). Women of Class 3 had an accelerated pace of aging as indicated by DunedinPoAm, but not an older epigenetic age as measured by PhenoAge or GrimAge. The association was significant among women who had ever used female hormones (β=0.521; 95%CI 0.014-1.027). Women of Class 1 or 2 did not exhibit accelerated epigenetic aging. Women of Class 3 had higher mortality (HR=1.40, 95%CI 1.08-1.81), and 36.3% of the effect was mediated through accelerated DunedinPoAm. Findings suggest that women with reproductive profiles characterized by early menopause may have altered epigenetic aging trajectories. Pace of aging may be more sensitive to the impact of reproductive profile variations than the status of biological age as indicated by PhenoAge or GrimAge. Clinically monitoring the pace of biological aging among women with early menopause and an appropriate application of hormone replacement therapy may minimize the negative consequence of accelerated biological aging and reduce premature mortality.

## Introduction

Evolutionary theories of aging are centered around the principle that aging is a byproduct of evolution because natural selection weakens after organisms’ reproductive periods (1). Disposable soma theory, for example, stipulates that scarce resources will shift energy allocation from repair to reproductive activities before dying, resulting in premature senescence and aging (2).

The trade-off between reproduction and aging has been empirically observed and is of great clinical significance in female humans (3). Reproductive profiles in women can be assessed through multiple indicators, including age at menarche, gravidity (i.e., number of pregnancies regardless of outcome), parity (i.e., number of births), and age at menopause (4). Early menarche and menopause and increased parity have been independently associated with higher risk of cardiovascular disease (CVD) and all-cause mortality (5–11). Parity exhibited a U-shaped relationship with all-cause mortality where both nulliparity (no live births) and high parity were found to be associated with higher mortality with the lowest risk observed for women with 3-4 live births (12–14). In addition, higher parity or grand multiparity (≥5 live births) was associated with increased risks of multiple diseases including kidney cancer (15), Type 2 diabetes (16), central nervous system disorders (17), and periodontal disease (18). This increased risk may be mediated by increased risk of metabolic syndrome (19,20) and elevated inflammation and oxidative stress (21–23). On the other hand, the potential benefit of an optimal level of parity versus nulliparity may be related to post-selection bias or psychosocial factors. Nulliparous women may be more likely to have chronic health problems that prevent them from giving birth and they may receive less social support which contributes to higher mortality (24–26).

Biological aging is a process of gradual accumulation of molecular and cellular damage leading to functional decline, diseases, and eventually death (27). While aging-related morbidities and mortality are direct indicators of accelerated biological aging, it takes considerable time and effort to track individuals’ life trajectories. To facilitate studies of geroprotective interventions, biomarkers of aging, including telomere length, physiological composite scores, and omics-based aging clocks, are emerging as surrogate endpoints (27–32). Epigenetic aging clocks were developed based on DNA methylation (DNAm) levels and have been found to explain variance in aging-related physical decline and mortality above and beyond chronological age (33). First-generation epigenetic clocks, such as Horvath (34) and Hannum clocks (35), were trained with chronological age (36). Second-generation clocks including PhenoAge and GrimAge, were trained with mortality and clinical biomarker data and have demonstrated relationships with healthspan and lifespan (37,38). Considering that the pace of biological aging may matter more than the a single measure of biological age, newer epigenetic clocks, including DunedinPoAm and DunedinPACE, have been trained to predict the pace of biological aging using within-individual data on clinical biomarkers longitudinally collected across two decades of follow-up (39,40).

Accelerated biological aging has been associated with increased reproductive effort and altered reproductive lifespan. For instance, higher gravidity was associated with shorter telomere length and accelerated epigenetic aging both cross-sectionally and longitudinally among young and early-middle-aged Filipino women (41,42). Increased parity was also associated with older epigenetic age in middle-aged and older non-Hispanic White females (43). Among post-menopausal women, a U-shaped pattern was observed for the relationship between parity and physiological age, with the youngest physiological age occurring in women with 3-4 live births (44). Beyond parity, evidence suggests that a shortened reproductive lifespan due to early menopause may also impact epigenetic aging. In middle-aged and older women, earlier age at menopause was associated with epigenetic age acceleration measured by Horvath and GrimAge clocks. This relationship may be related to hormone deficiency after menopause (38,45), as hormone replacement therapy (HRT) was associated with younger epigenetic age and reduced risk of mortality (45,46).

Though gravidity, parity, and reproductive lifespan have been analyzed independently as factors related to accelerated biological aging and future health risks, these factors are interconnected and should be considered jointly. Comprehensive reproductive profiles, accounting for gravidity, parity, and reproductive lifespan, remain to be explored in the context of biological aging and mortality. Using a nationally representative sample from the National Health and Nutrition Examination Survey (NHANES), the current study aimed to identify latent reproductive profiles among post-menopausal women and evaluate their associations with epigenetic aging. Further, we associated reproductive profiles with mortality data and examined the mediating effect of epigenetic aging. We hypothesized that women who paid excessive reproductive effort via higher gravidity or parity and had a shifted reproductive period, compared to the average, would exhibit accelerated epigenetic aging and higher mortality, with epigenetic aging mediating the impact of reproductive profiles on mortality.

## Methods

### Study design and data source

NHANES is a nationwide study conducted by the Centers for Disease Control and Prevention. It collected data on participant samples representative of the U.S. population. Survey and laboratory data collected from female participants in the 1999-2000 and 2001-2002 cycles were included. Mortality data were collected until 12/31/2019. The flow chart of participant selection can be found in **Supplementary eFigure 1**.

### Reproductive variables

Data on reproductive variables were collected through self-reported questionnaires. Age at menarche was defined as age when first menstrual period occurred. Age at menopause was calculated by adding 1 year to the age at last menstrual period for women who had irregular periods because of going/having gone through menopause. Gravidity was based on total number of reported pregnancies including live births, miscarriages, stillbirths, tubal pregnancies or abortions. Parity was defined as the total number of live births. Women who had missing data were excluded.

### DNAm-based variables

DNAm in whole blood was measured using the Illumina Infinium MethylationEPIC BeadChip v1.0. Data were processed through steps including color correction and background subtraction, outlier sample removal, imputation, normalization, and sample mismatch identification and removal. Details about the steps can be found in the data documentation (47). Pre-calculated epigenetic clocks including PhenoAge, GrimAge2 (denoted as GrimAge afterwards), and DunedinPoAm (Dunedin Pace of Aging) were used for current analyses. Biological sex and blood cell proportions were estimated based on DNAm. Women who had missing epigenetic clock data or inconsistent self-reported sex and DNAm predicted sex were excluded.

### Covariates

Women aged 85 years and older were excluded due to survivorship bias. Self-reported race/ethnicity was grouped into non-Hispanic White, non-Hispanic Black, Hispanic, and other. Federal Income-to-poverty ratio (IPR) ranged from 0 to 5. IPR greater than 5 was coded as 5. IPR was categorized into low income (IPR<1), middle income (1≤IPR<4), and high income (IPR≥4). Considering the relative high percentage (∼10%) of missing data in IPR, an additional level indicating missing income was created to accommodate the missingness while retaining statistical power. Education was dichotomized as below high school and high school or above. Body mass index (BMI) was categorized into healthy weight (BMI<25), overweight (25≤BMI<30), and obesity (BMI≥30). Analyses also used surgical histories on the reproductive system including hysterectomy (removal of uterus), and oophorectomy (removal of at least one of the ovaries). HRT history was defined as usage of female hormones including estrogen or progesterone in any forms but not for birth control or infertility.

### Statistical analyses

Epigenetic clock estimates for PhenoAge and GrimAge were residualized for chronological age. DunedinPoAm was not residulized for chronological age because this clock estimates pace of aging and is not correlated with chronological age. PhenoAge and GrimAge deviations and DunedinPoAm were further residualized for DNAm estimated blood cell proportions and z-standardized. The z-standardized clock deviations were used in the modeling analyses. A latent profile analysis (LPA) was conducted to identify latent classes of reproductive profiles based on age at menarche, gravidity, parity, and age at menopause. Associations between LPA class membership and epigenetic aging were examined using general linear regression. Pairwise comparisons between classes were conducted with Tukey’s Honestly Significant Difference tests. Associations of LPA class membership or epigenetic aging with mortality were examined using Cox proportional hazards models. Mediation of LPA class membership on mortality through epigenetic aging was also examined. As mortality was relatively common in the sample (46% deceased during the follow-up), a parametric survival regression model was used as part of the mediation analysis with an assumption that the time-to-event variable followed a Weibull distribution (48). Effects estimation and statistical testing for the mediation analysis were based on 1000 Monte Carlo simulations for bootstrap. Analyses were conducted using R 4.3.2 and accounted for the complex, multistage, and probability sampling design of NHANES.

## Results

### Sample and variable description

In total, 770 women aged 50 to 84.9 years (mean 65.0, SE 0.5) were included in the analyses. Weighted race composition includes 77.9% non-Hispanic White, 8.5% non-Hispanic Black, 9.4% Hispanic, and 4.2% other race/ethnicity. Most participants had middle income (46.2%) and at least high school education (71.9%). Mean age at menarche and menopause were 12.8 (SE 0.1) and 46.4 (SE 0.4) years, respectively. Mean gravidity and parity were 3.7 (SE 0.1) and 3.0 (SE 0.1), respectively. Variable distributions can be found in **Supplementary eTable 1** and **Supplementary eFigure 2**. The original estimates of PhenoAge and GrimAge were strongly correlated with chronological age (r=0.759 and 0.800), whereas DunedinPoAm was not (r=0.059, **Supplementary eFigure 3**).

### LPA with reproductive variables

Four latent classes were identified with an entropy of 0.81 (**Supplementary eTable 2**). Women of Class 1 (unweighted N=39, 5.1% of sample) had higher gravidity but average parity. Women of Class 2 (N=97, 12.6% of sample) had both higher gravidity and higher parity. Women of Class 3 (N=143, 18.6% of sample) had a younger age at menopause. Women of Class 4 (N=491, 63.8% of sample) were average in all the four reproductive variables (**Figure 1**). Descriptive analyses indicated that Class 2 had more women self-reporting non-Hispanic Black race/ethnicity, with lower income, without high school education, or with obesity. Classes 1 and 2 had a higher proportion of women self-reporting Hispanic ethnicity. Class 3 had the highest proportion of women with hysterectomy, oophorectomy or HRT history, and Class 2 had the lowest proportion of women who ever had HRT (**Table 1** for weighted and **Supplementary eTable 3** for unweighted statistics).

**Figure 1.**
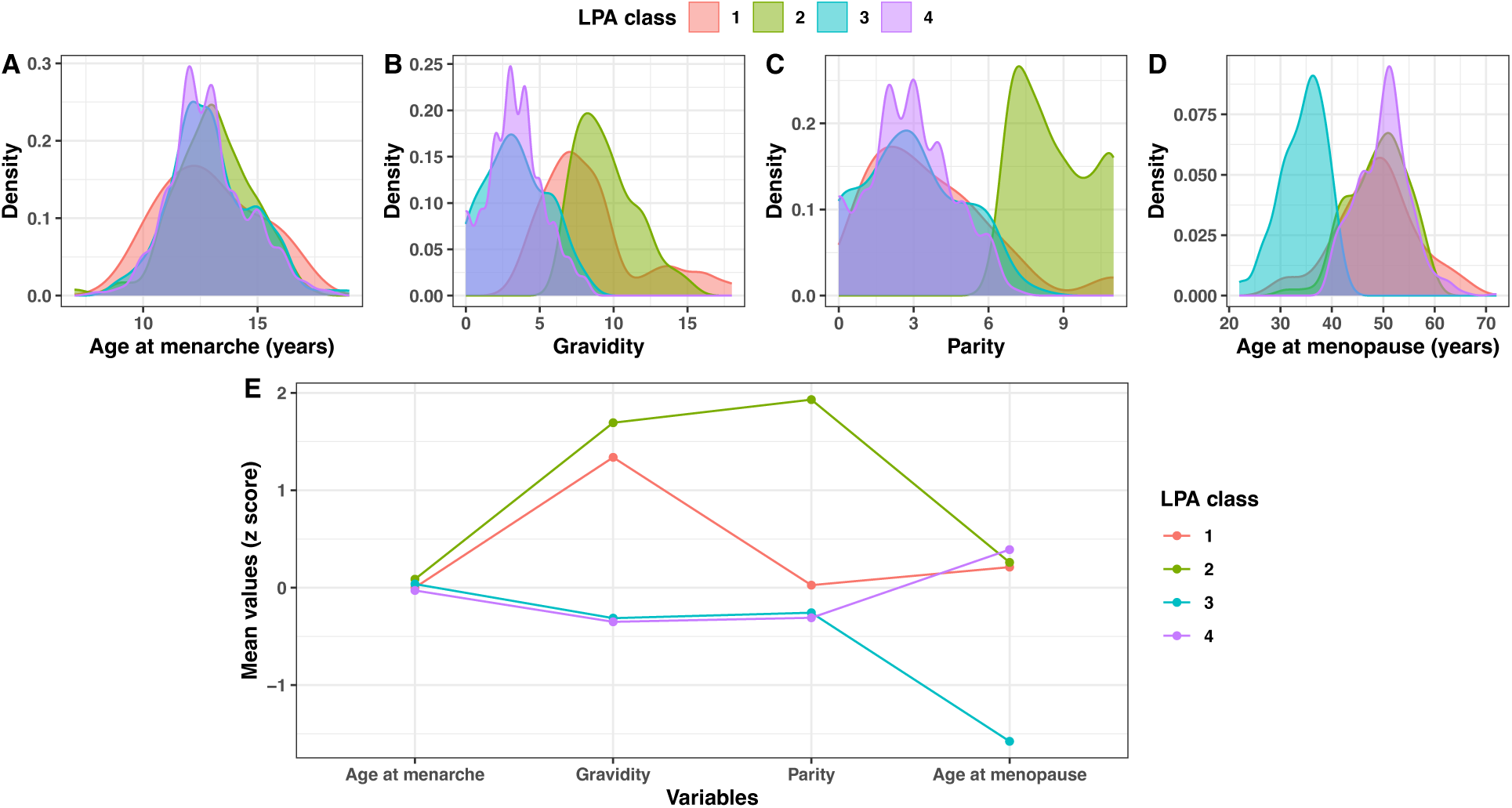
Characteristics of each of the four LPA classes. (**A-D**) Density plots (unweighted) showing the distribution of age at menarche, gravidity, parity, and age at menopause in each of the 4 classes identified by LPA. (**E**) Line plots showing the mean z scores (unweighted) of reproductive variables in each of the four classes identified by LPA.

**Table 1.**
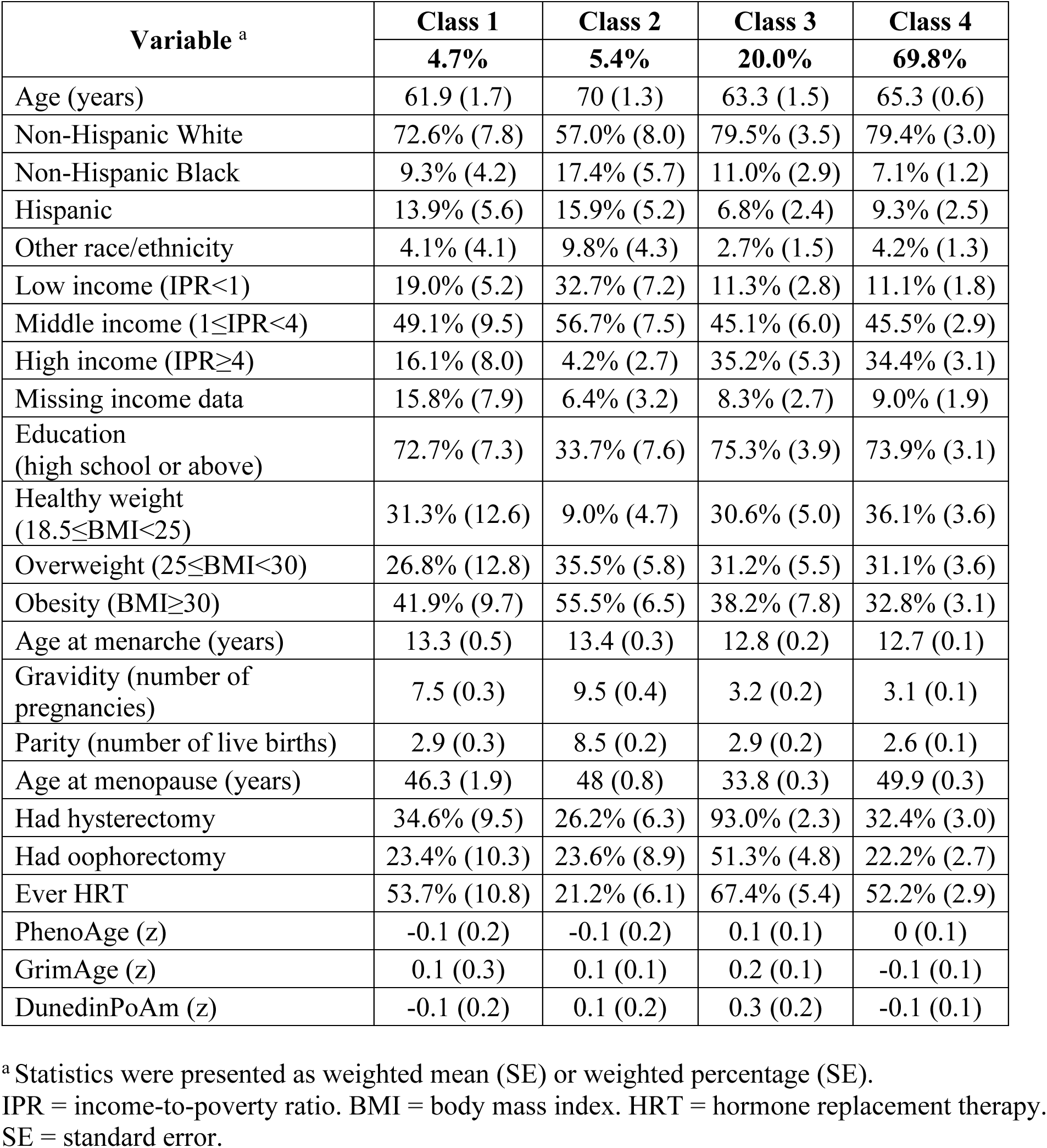
Weighted summary statistics of variables in each of the four latent classes.

### Latent class membership was associated with epigenetic aging

Pairwise comparison indicated that Class 3 (early menopause) had significantly accelerated DunedinPoAm (β=0.386, 95%CI 0.010-0.761, *P*=0.042) compared to Class 4 (average profile). PhenoAge and GrimAge did not show any significant differences between classes. Classes 1, 2, and 3 were not significantly different from each other in DunedinPoAm, but Class 3 had a higher mean epigenetic clock estimate than all other classes (**Figure 2A-C**). After adjusting for race/ethnicity, income, education, BMI, and hysterectomy, oophorectomy, and HRT histories the association was attenuated (Class 3 vs. Class 4 in DunedinPoAm: β=0.389, 95%CI −0.017 to 0.795, *P*=0.062) (**Figure 2D-F**).

**Figure 2.**
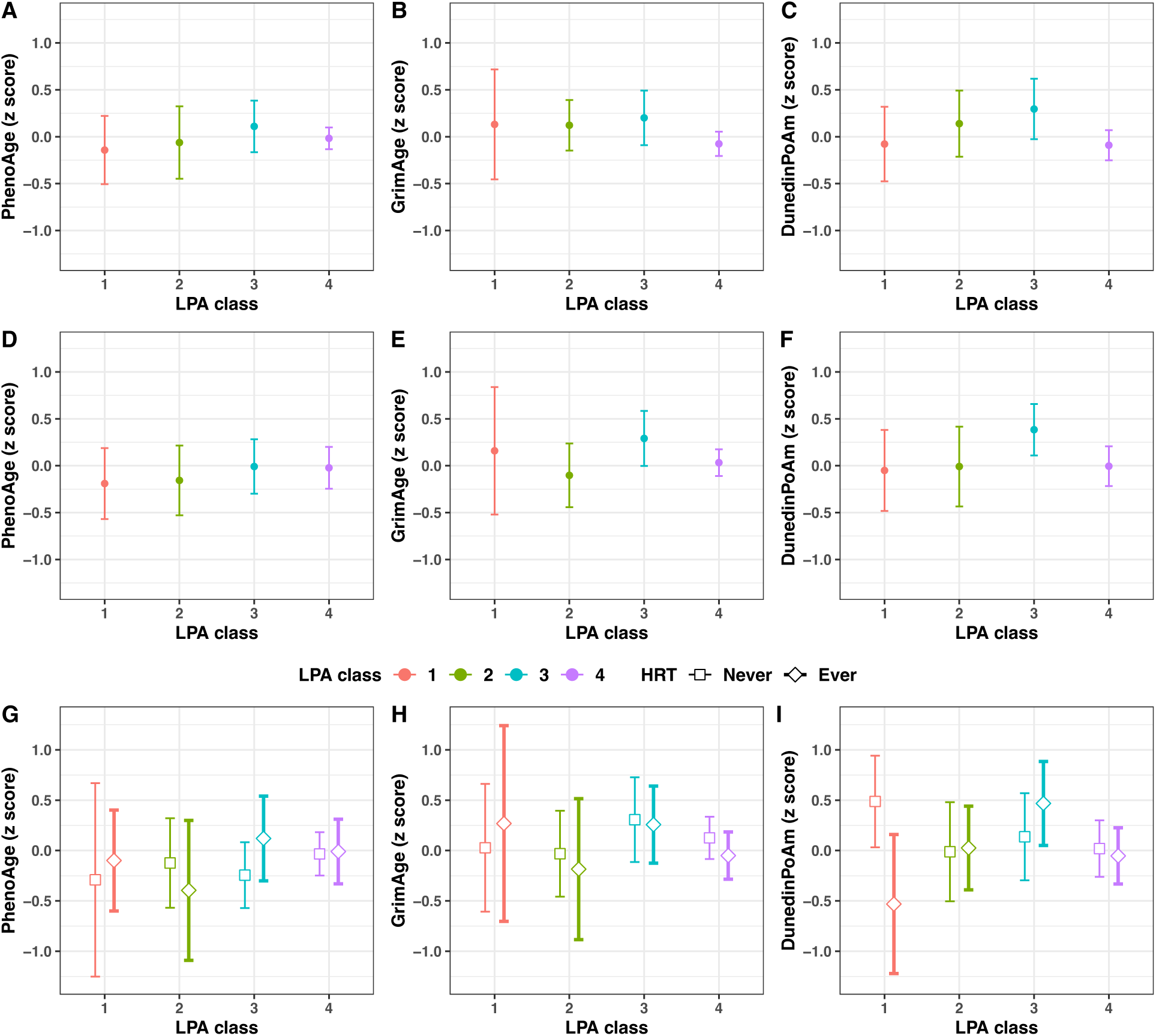
Epigenetic clock estimates (residualized for chronological age [not for DunedinPoAm] and cell proportions and z-standardized) (**A-C**) by LPA class without adjustment for covariates; (**D-F**) by LPA class with adjustment for race/ethnicity, income, education, BMI, hysterectomy, oophorectomy, and HRT history; and (**G-I**) by LPA class and HRT history after adjusting for race/ethnicity, income, education, BMI, and hysterectomy and oophorectomy. Analyses accounted for sampling design. LPA = latent profile analysis. HRT = hormone replacement therapy. BMI = body mass index.

Linear regression models including age at menarche, gravidity, parity, and age at menopause as individual predictors with epigenetic clocks as outcome variables revealed similar but not statistically significant trends (**Supplementary eFigure 4**).

### Latent class membership was associated with all-cause mortality

Among the 770 women, 351 were reported deceased during follow-up (**Supplementary eFigure 5**). Survival analyses indicated that women of Class 3 (early menopause) had higher all-cause mortality than women of Class 1, 2 or 4 while controlling for chronological age (HR=1.51, 95%CI 1.14-1.99) (**Supplementary eFigure 6**). Further adjustment for covariates including race/ethnicity, education, income, BMI, hysterectomy, oophorectomy, and HRT attenuated the relationship, but it remained significant (HR=1.40, 95%CI 1.08-1.81, **Table 2**). However, analyses with the top two specific causes of mortality – heart disease-related or malignant neoplasm-related mortality – showed no significant impact of Class 3 membership (**Supplementary eTable 4**).

**Table 2.** Cox proportional hazards regression models ^a^ predicting all-cause mortality.

| Predictor | Model 1 |  | Model 2 |  | Model 3 |  |
| --- | --- | --- | --- | --- | --- | --- |
|  | HR | 95% CI | HR | 95% CI | HR | 95% CI |
| LPA Class3 vs. Class 1,2,4 | **1.51** | 1.14 to 1.99 | **1.52** | 1.16 to 2.00 | **1.40** | 1.08 to 1.81 |
| Age>65 vs. Age≤65 | **5.48** | 3.83 to 7.83 | **4.82** | 3.35 to 6.93 | **4.57** | 3.18 to 6.57 |
| NH black vs. NH white |  |  | 0.87 | 0.63 to 1.20 | 0.79 | 0.54 to 1.14 |
| Hispanic vs. NH white |  |  | 0.73 | 0.47 to 1.15 | 0.71 | 0.44 to 1.14 |
| Other race/ethnicity vs. NH white |  |  | 1.04 | 0.48 to 2.26 | 1.01 | 0.46 to 2.23 |
| HS and above vs. Below HS |  |  | 0.87 | 0.63 to 1.20 | 0.90 | 0.66 to 1.25 |
| Low income vs. Middle income |  |  | 1.35 | 0.94 to 1.94 | 1.31 | 0.93 to 1.86 |
| High income vs. Middle income |  |  | **0.61** | 0.44 to 0.84 | **0.64** | 0.48 to 0.86 |
| Missing income vs. Middle income |  |  | 0.80 | 0.52 to 1.21 | 0.86 | 0.56 to 1.30 |
| Overweight vs. Healthy weight |  |  | 0.96 | 0.73 to 1.26 | 0.95 | 0.72 to 1.24 |
| Obese vs. Healthy weight |  |  | 0.98 | 0.75 to 1.29 | 0.99 | 0.76 to 1.28 |
| Had hysterectomy vs. No |  |  |  |  | **1.34** | 1.01 to 1.78 |
| Had oophorectomy vs. No |  |  |  |  | 1.06 | 0.75 to 1.51 |
| Ever HRT vs. No |  |  |  |  | **0.69** | 0.48 to 1.00 |

<sup>a</sup> Analyses accounted for sampling design.
LPA = latent profile analysis. NH = non-Hispanic. HS = high school. HRT = hormone replacement therapy. HR = hazard ratio.

All three epigenetic clocks significantly predicted all-cause mortality both before and after covariate adjustment (PhenoAge HR=1.25, 95%CI 1.06-1.47; GrimAge HR=1.59, 95%CI 1.35-1.86; DunedinPoAm HR=1.42, 95%CI 1.22-1.65; based on adjusted models; **Supplementary eFigure 7**).

The association between class membership and mortality was mediated by DunedinPoAm, but not PhenoAge or GrimAge (**Table 3**). Class 3 membership was associated with 26.50 (95% CI: 7.21-47.62) months shorter survival time as compared with Class 1, 2, or 4, the effects of which were mediated by increased DunedinPoAm, accounting for 36.3% of the total effects.

**Table 3.** Mediation of the association between LPA class membership and mortality through epigenetic aging.

| Mediator <sup>a</sup><br>Effect type <sup>b</sup> | PhenoAge |  |  | GrimAge |  |  | DunedinPoAm |  |  |
| --- | --- | --- | --- | --- | --- | --- | --- | --- | --- |
|  | Estimate | 95%CI | P value | Estimate | 95%CI | P value | Estimate | 95%CI | P value |
| **Average causal mediation effect** | −1.18 | −13.51 to 11.98 | 0.860 | −20.37 | −47.97 to 5.09 | 0.114 | −26.50 | −47.62 to −7.21 | **0.006** |
| **Average direct effect** | −67.24 | −127.26 to −9.77 | **0.028** | −65.14 | −127.89 to −8.82 | **0.032** | −44.99 | −105.74 to 14.42 | 0.136 |
| **Proportion mediated** | 2.06% | −32.91%<br>to 34.26% | 0.856 | 23.37% | −8.37%<br>to 72.70% | 0.118 | 36.31% | 9.01%<br>to 179.09% | **0.022** |

<sup>a</sup> General linear regression models were used as mediator models with mediators being included as continuous variables. Parametric survival regression models were used as outcome models with an assumption that the time-to-event variable followed a Weibull distribution. Mediators were z-scored estimates of epigenetic clocks.
<sup>b</sup> Effects estimation and statistical testing were based on 1000 Monte Carlo simulations for bootstrap. Analyses accounted for sampling design and were controlled for age, race/ethnicity, income level, BMI category, hysterectomy and oophorectomy history, and history of hormone replacement therapy.

### Interaction between HRT and reproductive profiles

The association between reproductive profiles and DunedinPoAm was significantly moderated by HRT history (*χ*^2^(3)=10.61, *P*=0.014). Class 3 was significantly different from Class 4 in DunedinPoAm only among women who had ever used female hormones (β=0.521, 95%CI 0.014-1.027, *P*=0.043, **Figure 2G-I**). Among women in Class 1, those who had used female hormones had significantly lower DunedinPoAm than those who had not (β=−1.018, 95%CI −1.875 to −0.160, *P*=0.024), indicating slower pace of aging. However, no interaction between HRT and reproductive profiles was found with all-cause mortality.

Post-hoc analyses with specific HRT details revealed that women of Class 1 were more likely to still have menstrual cycles when starting HRT (54% vs. 4%-25% in other classes), whereas women of Class 3 were more likely to have started HRT at a younger age (43 vs. 51 years old in other classes) with a longer duration of HRT (13 vs. 6-8 years in other classes) (**Supplementary eTable 5**).

## Discussion

Drawing on a nationally representative sample of post-menopausal women, we identified four latent reproductive profiles and found that women with early menopause had accelerated pace of epigenetic aging. HRT history significantly moderated the association, such that women with early menopause had accelerated DunedinPoAm only when they ever had HRT. Women with early menopause were also found to have higher all-cause mortality, mediated by accelerated pace of aging.

The health benefit of female HRT has a controversial history (49). Estrogen-only products were first approved for the treatment of hot flashes induced by menopause, but were later associated with increased risk of endometrial cancer. Combination therapy with both estrogen and progesterone was found to be beneficial in reducing the risk of endometrial cancer, CVD, osteoporosis, dementia, and all-cause mortality (50). However, clinical trials revealed potential risks including coronary heart disease, stroke, pulmonary embolism, and breast cancer (50). To maximize the benefit-to-risk ratio, current guidelines recommend that HRT should be used for women aged ≤60 years, initiated within 10 years of menopause onset, and used for the shortest total duration and with the lowest effective dose (50). Similarly, we found contradictory effects of HRT for women with different reproductive profiles. For women with high gravidity and average parity, HRT was found to be associated with decelerated epigenetic aging pace. In contrast, for women with early menopause, HRT might be associated with accelerated epigenetic aging pace although it was not significant. The effect difference could be related to initiation timing, duration, dosage, and route of administration of HRT tailored for women with different reproductive profiles. It is also possible that women who underwent HRT were more symptomatic which might be related to accelerated aging pace. Nevertheless, analyses with mortality as the outcome replicated previous findings that HRT was associated with lower all-cause mortality. No interaction was found between reproductive profiles and HRT in predicting all-cause mortality.

The association between reproductive profiles and all-cause mortality is consistent with prior research. A previous study using data from NHANES 2003-2018 also found that later menopause was associated with decreased all-cause mortality (7). A meta-analysis of 310,329 women across 32 studies reported a significantly higher CVD mortality in women with an age at menopause<45 years (9), though our study did not find a significant association with cause-specific mortality. This may be related to insufficient power due to relatively small effect size and sample size. Although age at menarche is considered as a reproductive milestone, it did not contribute substantially to the classification of reproductive profiles as compared with gravidity, parity, and age at menopause. This could be due to limited variations in age at menarche and the effect dilution of early-life milestones by accumulated impact of other life experiences as women age. Further, the association between reproductive profiles and all-cause mortality was significantly mediated through accelerated DunedinPoAm, but not through PhenoAge or GrimAge, suggesting that pace of aging may be more sensitive to the impact of reproductive variations than status of biological age. Nevertheless, the association between reproductive profiles and mortality was the greatest for GrimAge, followed by DunedinPoAm and PhenoAge, which is consistent with the ranking of effect sizes among the three clocks as reported in previous studies (37–40).

Inconsistent with evolutionary theories of aging, reproductive profiles characterized by high gravidity or parity were not associated with epigenetic aging acceleration. Considering a potential U-shaped pattern (44), post-hoc analyses were conducted by including a quadratic term of gravidity or parity in regression models. The results did not support a significantly non-linear relationship. As suggested in previous studies, the effect size of the relationship was relatively small, and would be attenuated further after adjusting for covariates (43). Our sample size may not be large enough to detect these effects. Nevertheless, as suggested by the results with DunedinPoAm, women with high gravidity and without a history of HRT might have a trend of accelerated epigenetic aging as compared with women with an average reproductive profile.

### Strengths and limitations

The strengths of this study include a nationally representative sample, making our results generalizable to the population across the U.S. The use of LPA technique with a set of reproductive health variables facilitated the identification of latent reproductive profiles. In addition, the presence of both epigenetic clock and mortality data in the dataset enabled mediation analyses, which had not been conducted in previous studies.

We also acknowledge limitations. First, the epigenetic clock data in NHANES was only available for years 1999-2002 and for participants aged ≥50 years. Future data release for other cycles of survey and for younger participants would enable studies of more comprehensive reproductive profiles and boost statistical power. Second, DunedinPoAm was later updated to DunedinPACE which showed improved performance (40). However, DunedinPACE was not available from the publicly available NHANES data repository. Future studies should seek to replicate results with DunedinPACE. Third, many variables in NHANES, including age at menarche, were self-reported, which introduces recall bias. This is particularly salient given the long-time span between the onset of menarche and the participants reaching their 50th birthday, which may affect the accuracy of these data. Finally, constrained by sample size, interactions between detailed HRT history and reproductive profile were not examined. Future studies in larger, more diverse samples should include detailed HRT data and stratified analyses to better understand these interactions.

## Conclusions

Four reproductive profiles were identified among post-menopausal women. Women with early menopause exhibited accelerated pace of epigenetic aging, the effect of which was pronounced among women who had ever received HRT. Inconsistent with evolutionary theories of aging, women who had high gravidity or parity did not show accelerated epigenetic aging. The reproductive profile characterized by early menopause was significantly associated with higher mortality, and about a third of the effect was mediated through accelerated pace of epigenetic aging. DunedinPoAm may be more sensitive in reflecting the impact of reproductive profiles on aging trajectories than PhenoAge or GrimAge. Findings also suggest that age at menopause may be a prominent factor in shaping epigenetic aging trajectories. Monitoring the pace of biological aging for women with early menopause and applying an appropriate regimen of hormone therapy may minimize the negative consequence of accelerated biological aging and reduce premature mortality.

## Data Availability

All the data used in this study are publicly downloadable from the website of National Center for Health Statistics (https://www.cdc.gov/nchs/nhanes/index.html).

## Author contribution

Q.Y., L.E., and I.S conceptualized the study. Q.Y. conducted data analyses and drafted the manuscript. A.A., I.S. and L.E. reviewed and approved the manuscript.

