## Supplementary Material for "Evaluation of reproductive profiles, epigenetic aging, and mortality in post-menopausal women"

**Supplementary Materials for**
**Evaluation of reproductive profiles, epigenetic aging, and mortality in post-menopausal women**

Qiaofeng Ye, MS[1]; Aaliya Ahamed, BS[1]; Idan Shalev, PhD[1]; Laura Etzel, PhD[2*]

**This document includes:**

**eFigure 1. Flow chart of participant selection.**

**eFigure 2. Distribution of continuous variables.**

**eFigure 3. Density and scatter plots of chronological age and epigenetic clocks.**

**eFigure 4. Effect size estimates from linear regression including age at menarche, gravidity, parity, and age at menopause as individual predictors, and epigenetic clocks as outcomes.**

**eFigure 5. Causes of mortality.**

**eFigure 6. Kaplan-Meier survival curve (all-cause mortality) by LPA class and age category.**

**eFigure 7. Associations between epigenetic aging and mortality.**

**eTable 1. Descriptive statistics of demographic and reproductive variables.**

**eTable 2. Model selection process of latent profile analysis.**

**eTable 3. Unweighted descriptive statistics of variables in each of the four latent classes.**

**eTable 4. Cox proportional hazards regression models predicting heart disease-relate mortality and malignant neoplasm-related mortality.**

**eTable 5. Detailed information about HRT among women who ever had HRT by latent reproductive profile.**

### Supplemental Figures

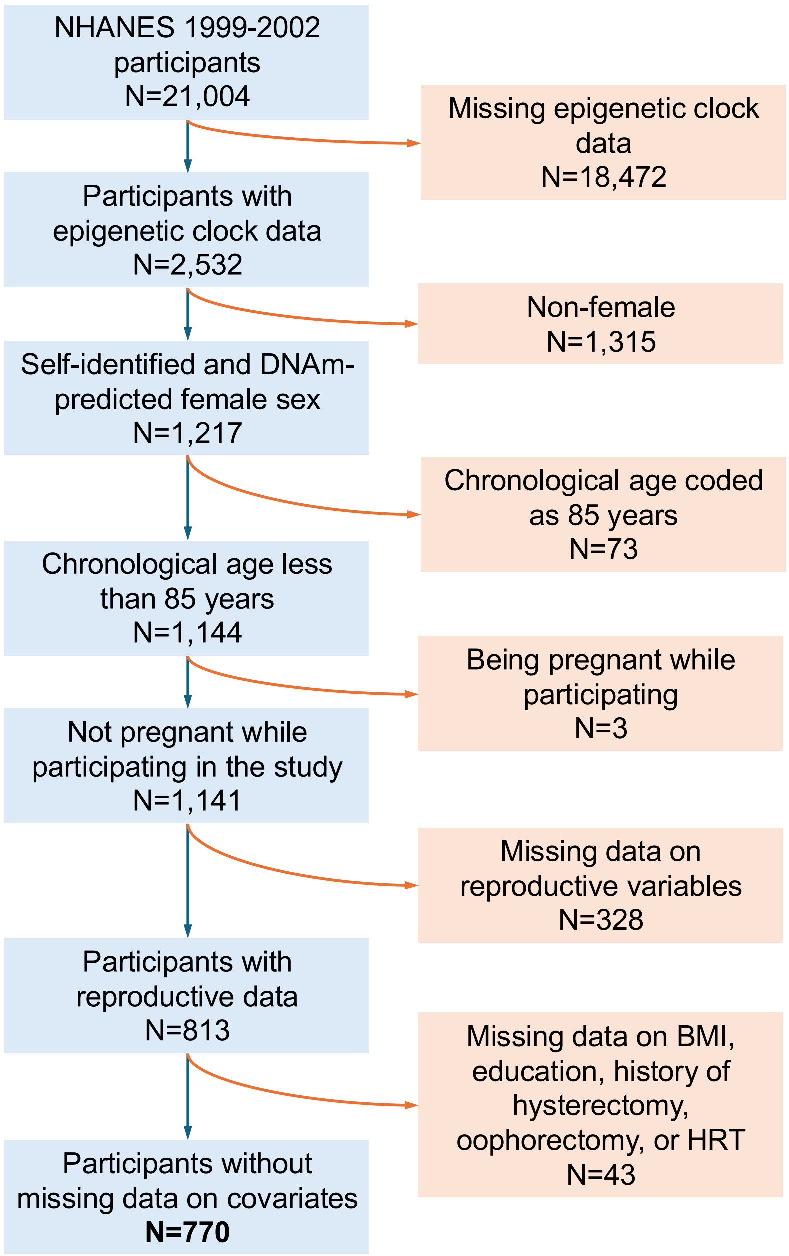


**eFigure 1. Flow chart of participant selection.**
DNAm = DNA methylation. BMI = body mass index. HRT = hormone replacement therapy.

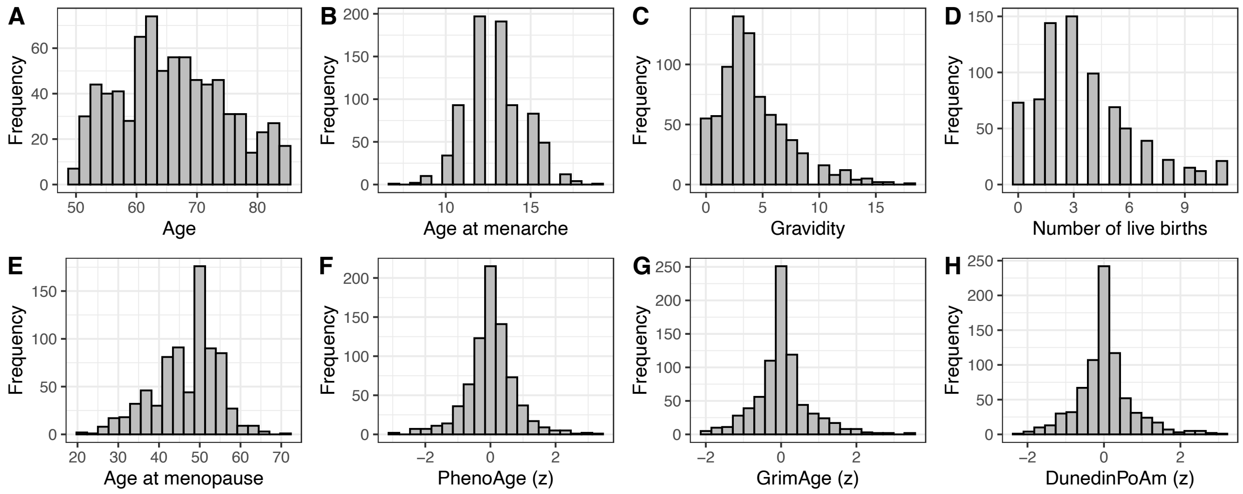


**eFigure 2. Distribution of continuous variables.**

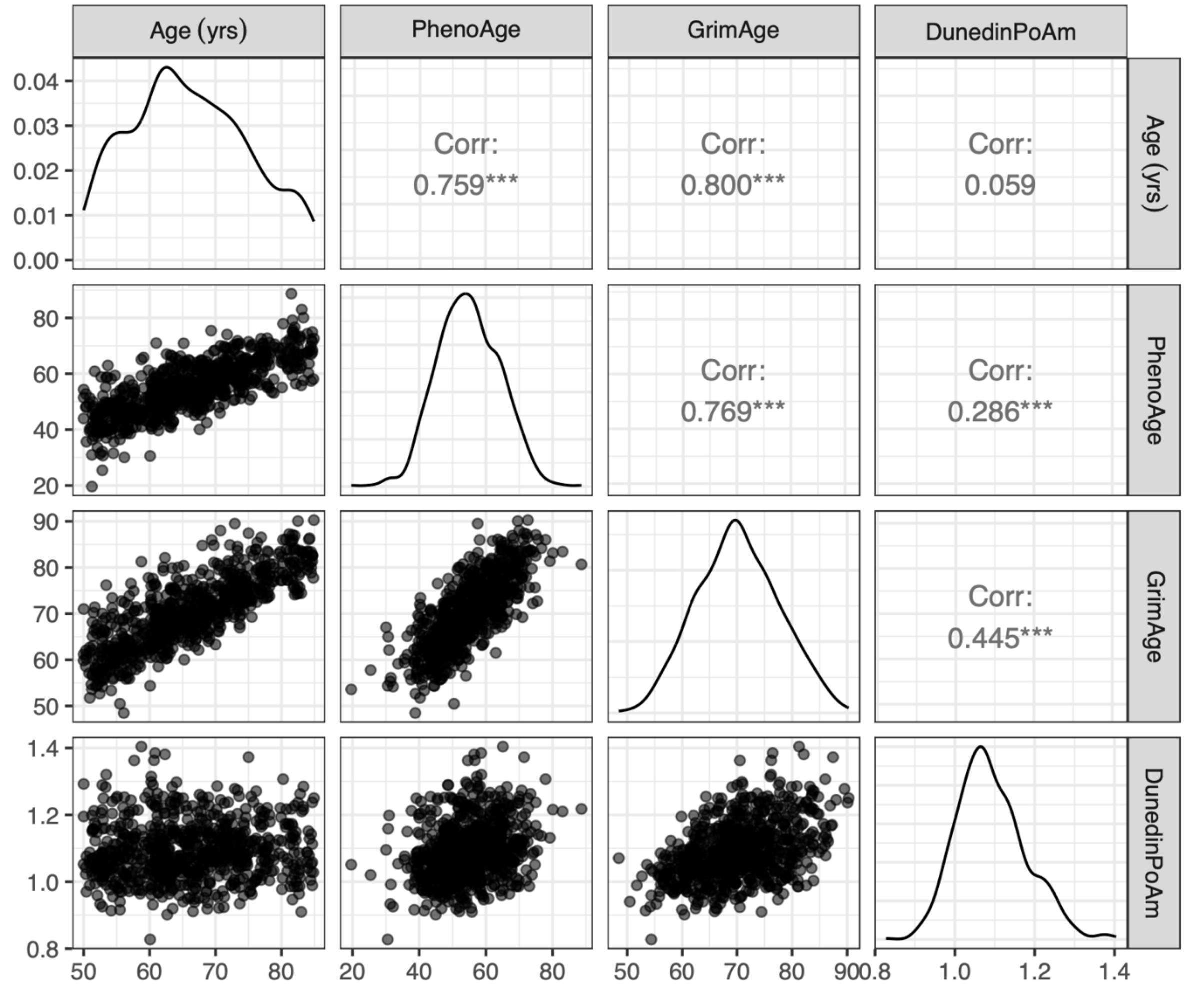


**eFigure 3. Density and scatter plots of chronological age and epigenetic clocks.**
The epigenetic clock values in the plots are original estimated without any residualization or standardization. Pearson's correlation coefficients are printed on the upper triangular panels.

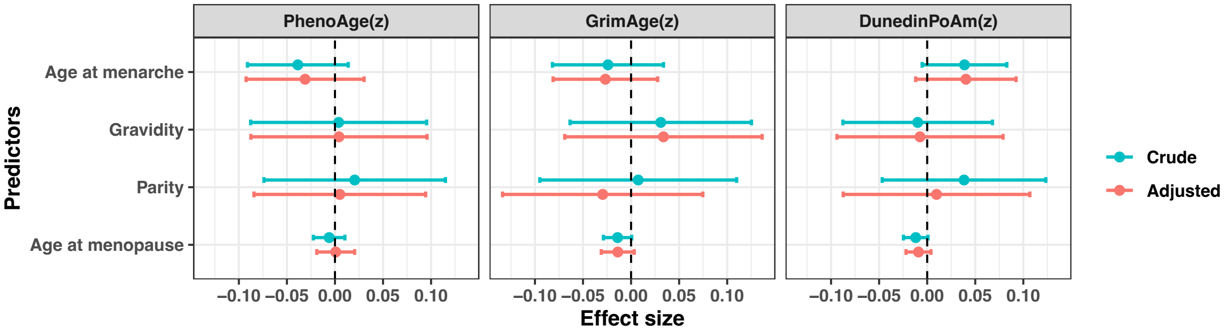


**eFigure 4. Effect size estimates from linear regression including age at menarche, gravidity, parity, and age at menopause as individual predictors, and epigenetic clocks as outcomes.**

The crude models only included the four predictors. The adjusted models controlled for covariates including race/ethnicity, education, income, body mass index, and histories of hysterectomy, oophorectomy, and hormone replacement therapy. Analyses accounted for sampling design.

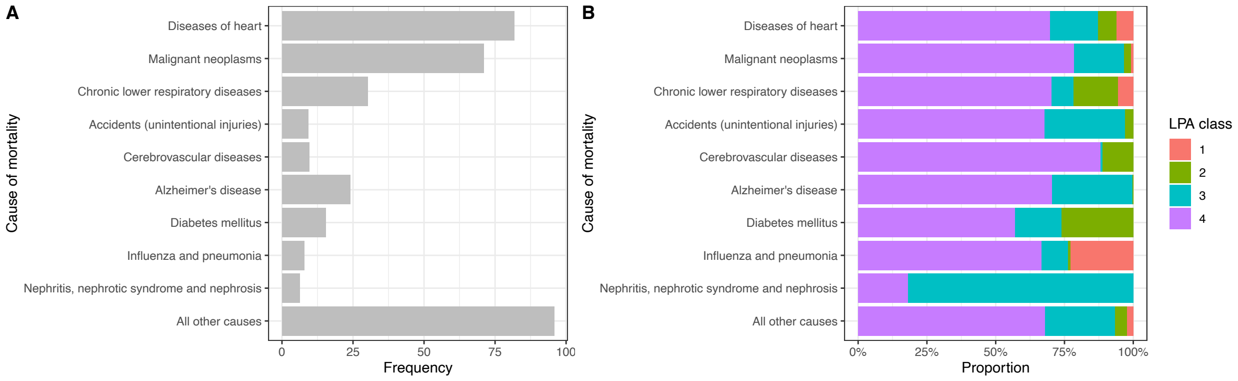


### eFigure 5. Causes of mortality.

(**A**) Weighted frequencies of causes of mortality. (**B**) Weighted proportions of participants belonging to each LPA class for each cause of mortality.

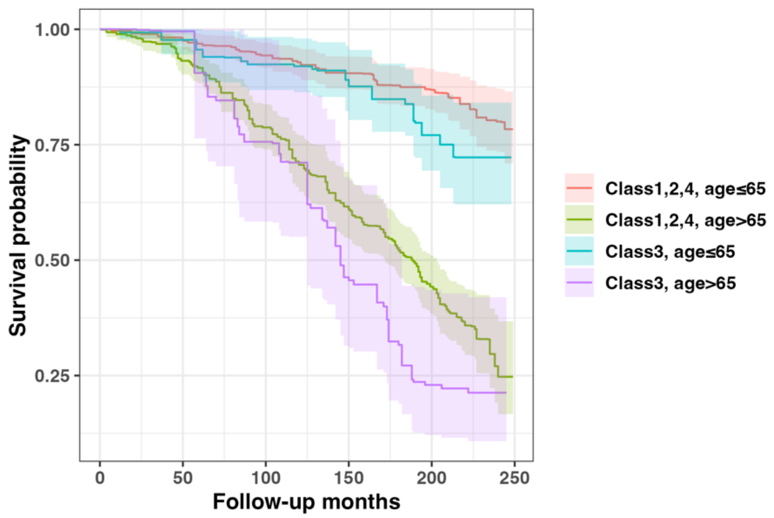


**eFigure 6. Kaplan-Meier survival curve (all-cause mortality) by LPA class and age category.**
Analyses accounted for sampling design.

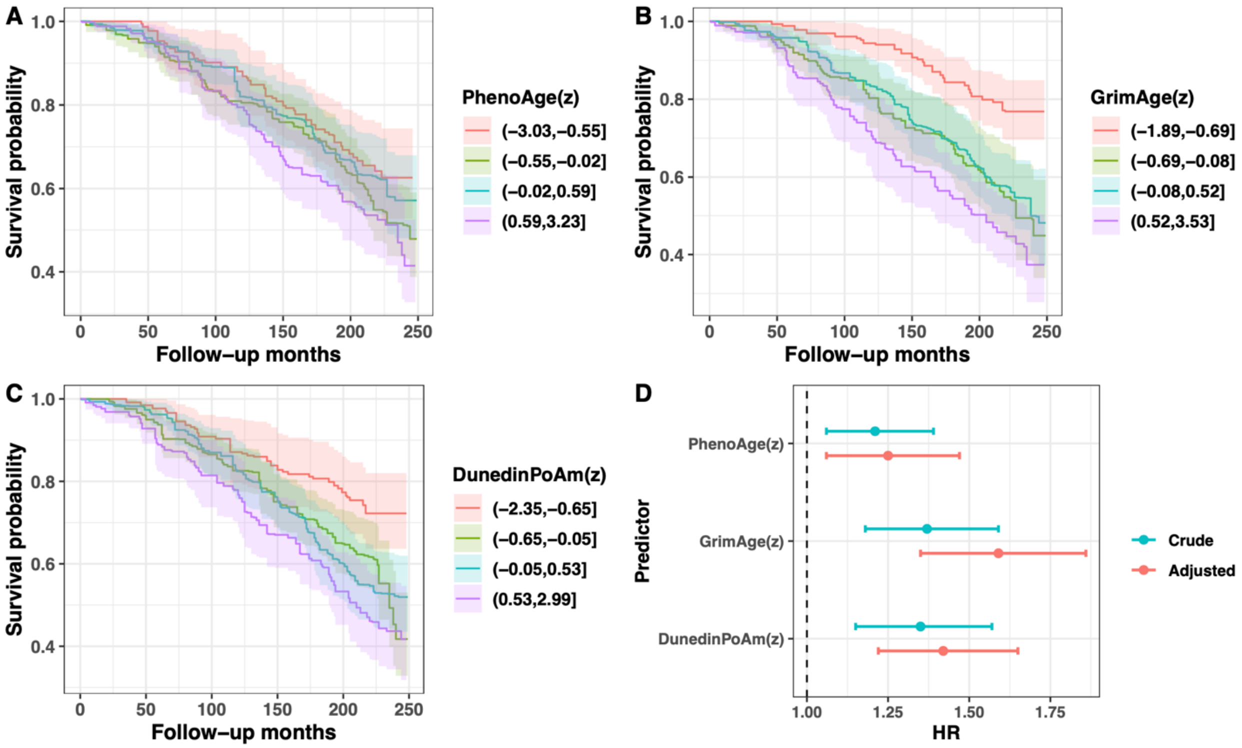


### eFigure 7. Associations between epigenetic aging and mortality.

(**A-C**) Kaplan-Meier survival curves (all-cause mortality) by epigenetic clock quartile (residualized for chronological age [not for DundedinPoAm] and cell proportion, and z-scored). (**D**) Forest plot showing hazard ratio by epigenetic clock. The three types of clock estimates (residualized for chronological age [not for DundedinPoAm] and cell proportion, and z-scored) were included as continuous predictors in separate Cox proportional hazard regression models. Crude models only included clock estimates as the predictor for mortality. Adjusted models were controlled for covariates including age, race/ethnicity, income, BMI, and histories of hysterectomy, oophorectomy and hormone replacement therapy. Analyses accounted for sampling design. HR = hazard ratio.

### Supplemental Tables

#### eTable 1. Descriptive statistics of demographic and reproductive variables.

| Variable | Mean (SD) or N (%) | Weighted mean (SE) or Weighted percentage (SE) | Range (min / max) |
|---|---|---|---|
| Total | 770 (100%) | | |
| Age (years) | 66.0 (8.8) | 65.0 (0.5) | 50.0 / 84.9 |
| Non-Hispanic White | 312 (40.5%) | 77.9% (2.8) | |
| Non-Hispanic Black | 159 (20.6%) | 8.5% (1.5) | |
| Hispanic | 268 (34.8%) | 9.4% (2.3) | |
| Other race/ethnicity | 31 (4.0%) | 4.2% (1.2) | |
| Low income (IPR<1) | 122 (15.8%) | 12.7% (1.7) | |
| Middle income (1≤IPR<4) | 391 (50.8%) | 46.2% (2.3) | |
| High income (IPR≥4) | 178 (23.1%) | 32.0% (2.6) | |
| Missing income data | 79 (10.3%) | 9.0% (1.7) | |
| Education (high school or above) | 432 (56.1%) | 71.9% (2.5) | |
| Healthy body weight (18.5≤BMI<25) | 199 (25.8%) | 33.3% (2.4) | |
| Overweight (25≤BMI<30) | 258 (33.5%) | 31.1% (2.9) | |
| Obesity (BMI≥30) | 313 (40.6%) | 35.6% (2.9) | |
| Age at menarche (years) | 12.9 (1.7) | 12.8 (0.1) | 7.0 / 19.0 |
| Gravidity (number of pregnancies) | 4.4 (3.0) | 3.7 (0.1) | 0 / 18.0 |
| Parity (number of live births) | 3.6 (2.6) | 3.0 (0.1) | 0 / 11.0 |
| Age at menopause (years) | 46.9 (7.9) | 46.4 (0.4) | 22.0 / 72.0 |
| Had hysterectomy | 317 (41.2%) | 44.3% (2.5) | |
| Had at least one ovary removed | 201 (26.1%) | 28.2% (2.4) | |
| Ever HRT | 331 (43.0%) | 53.6% (2.4) | |

IPR = income-to-poverty ratio. BMI = body mass index. HRT = hormone replacement therapy.

**eTable 2. Model selection process of latent profile analysis.**

| Model | Classes | AIC | BIC | Entropy | prob_min | prob_max | n_min | n_max | BLRT_p |
|---|---|---|---|---|---|---|---|---|---|
| EEE | 1 | 14603.11 | 14668.16 | 1 | 1 | 1 | 1 | 1 | |
| EEE | 2 | 14567.06 | 14655.35 | 0.66 | 0.8 | 0.95 | 0.26 | 0.74 | 0.01 |
| EEE | 3 | 14408.53 | 14520.05 | 0.77 | 0.82 | 0.94 | 0.14 | 0.66 | 0.01 |
| **EEE** | **4** | **14128.48** | **14263.23** | **0.81** | **0.81** | **0.93** | **0.05** | **0.64** | **0.01** |
| EEE | 5 | 14128.99 | 14286.97 | 0.79 | 0.59 | 0.95 | 0.05 | 0.65 | 0.1 |
| EEE | 6 | 14138.94 | 14320.15 | 0.58 | 0 | 0.96 | 0 | 0.62 | 0.12 |
| EEE | 7 | 14159.49 | 14363.93 | 0.48 | 0 | 0.96 | 0 | 0.3 | 0.95 |
| EEE | 8 | 14169.65 | 14397.32 | 0.44 | 0 | 0.96 | 0 | 0.56 | 0.78 |
| EEE | 9 | 14163.17 | 14414.08 | 0.47 | 0 | 0.97 | 0 | 0.55 | 0.06 |

EEE = equal volume, equal shape, and equal orientation

### eTable 3. Unweighted descriptive statistics of variables in each of the four latent classes.

| Variable [a] | Class 1 | Class 2 | Class 3 | Class 4 |
|---|---|---|---|---|
| | N=39 | N=97 | N=143 | N=491 |
| Age (years) | 62.0 (7.7) | 68.1 (7.5) | 64.2 (8.4) | 66.5 (9.1) |
| Non-Hispanic White | 13 (33.3%) | 15 (15.5%) | 58 (40.6%) | 226 (46.0%) |
| Non-Hispanic Black | 7 (17.9%) | 20 (20.6%) | 39 (27.3%) | 93 (18.9%) |
| Hispanic | 18 (46.2%) | 58 (59.8%) | 42 (29.4%) | 150 (30.5%) |
| Other race/ethnicity | 1 (2.6%) | 4 (4.1%) | 4 (2.8%) | 22 (4.5%) |
| Low income | 11 (28.2%) | 31 (32.0%) | 18 (12.6%) | 62 (12.6%) |
| Middle income | 17 (43.6%) | 48 (49.5%) | 74 (51.7%) | 252 (51.3%) |
| High income | 5 (12.8%) | 7 (7.2%) | 37 (25.9%) | 129 (26.3%) |
| Missing income data | 6 (15.4%) | 11 (11.3%) | 14 (9.8%) | 48 (9.8%) |
| Education (high school or above) | 19 (48.7%) | 18 (18.6%) | 84 (58.7%) | 311 (63.3%) |
| Healthy weight | 8 (20.5%) | 10 (10.3%) | 38 (26.6%) | 143 (29.1%) |
| Overweight | 13 (33.3%) | 36 (37.1%) | 46 (32.2%) | 163 (33.2%) |
| Obesity | 18 (46.2%) | 51 (52.6%) | 59 (41.3%) | 185 (37.7%) |
| Age at menarche (years) | 12.9 (2.1) | 13.1 (1.7) | 13.0 (1.8) | 12.9 (1.7) |
| Gravidity (number of pregnancies) | 8.4 (3.4) | 9.5 (2.0) | 3.4 (2.1) | 3.3 (1.9) |
| Parity (number of live births) | 3.7 (2.5) | 8.7 (1.5) | 3.0 (2.0) | 2.8 (1.7) |
| Age at menopause (years) | 48.6 (7.8) | 49.0 (5.7) | 34.4 (4.4) | 50.0 (5.1) |
| Had hysterectomy | 11 (28.2%) | 29 (29.9%) | 127 (88.8%) | 150 (30.5%) |
| Had oophorectomy | 7 (17.9%) | 19 (19.6%) | 74 (51.7%) | 101 (20.6%) |
| Ever HRT | 13 (33.3%) | 21 (21.6%) | 81 (56.6%) | 216 (44.0%) |
| PhenoAge (z) | -0.1 (0.8) | 0.04 (0.5) | 0.1 (0.7) | 0.01 (0.8) |
| GrimAge (z) | 0.03 (0.8) | 0.1 (0.4) | 0.1 (0.8) | -0.02 (0.7) |
| DunedinPoAm (z) | -0.03 (0.7) | 0.1 (0.4) | 0.2 (0.8) | -0.03 (0.7) |

[a] Statistics were not weighted, and presented as mean (SD) or N (%).
HRT = hormone replacement therapy.

**eTable 4. Cox proportional hazards regression models predicting heart disease-relate mortality and malignant neoplasm-related mortality.**

| | Heart diseases-related mortality | | Malignant neoplasm-related mortality | |
|---|---|---|---|---|
| **Predictor** | **HR** | **95% CI** | **HR** | **95% CI** |
| LPA Class3 vs. Class 1,2,4 | 1.15 | 0.63 to 2.11 | 1.40 | 0.75 to 2.60 |
| Age>65 vs. Age≤65 | **5.81** | 2.99 to 11.3 | **4.76** | 2.40 to 9.41 |
| NH black vs. NH white | 0.66 | 0.31 to 1.41 | 0.64 | 0.28 to 1.46 |
| Hispanic vs. NH white | 0.44 | 0.18 to 1.10 | 0.67 | 0.32 to 1.40 |
| Other race/ethnicity vs. NH white | 0.65 | 0.08 to 5.09 | 1.36 | 0.28 to 6.52 |
| HS and above vs. Below HS | 0.55 | 0.30 to 1.01 | 1.25 | 0.68 to 2.31 |
| Low income vs. Middle income | 1.62 | 0.78 to 3.36 | 1.29 | 0.46 to 3.64 |
| High income vs. Middle income | 0.48 | 0.20 to 1.12 | 0.68 | 0.29 to 1.60 |
| Missing income vs. Middle income | 0.45 | 0.10 to 2.04 | 1.34 | 0.49 to 3.64 |
| Overweight vs. Healthy weight | 1.37 | 0.65 to 2.92 | 1.02 | 0.50 to 2.11 |
| Obese vs. Healthy weight | 1.61 | 0.83 to 3.11 | 1.55 | 0.66 to 3.63 |
| Had hysterectomy vs. No | 1.64 | 0.66 to 4.07 | 0.93 | 0.45 to 1.89 |
| Had oophorectomy vs. No | 1.04 | 0.42 to 2.54 | 1.46 | 0.65 to 3.29 |
| Ever HRT vs. No | 0.85 | 0.43 to 1.69 | 0.58 | 0.23 to 1.49 |

LPA = latent profile analysis. NH = non-Hispanic. HS = high school. HRT = hormone replacement therapy. HR = hazard ratio. Analyses accounted for sampling design.

**eTable 5. Detailed information about HRT among women who ever had HRT by latent reproductive profile.**

| Variables | Class 1 | Class 2 | Class 3 | Class 4 |
|---|---|---|---|---|
| Still having periods when started HRT | 53.6% (14.4) | 3.6% (2.7) | 5.1% (3.5) | 25.0% (4.5) |
| Age at HRT initiation (years) | 50.6 (2.3) | 50.9 (1.6) | 43.0 (2.1) | 50.8 (0.7) |
| Duration of HRT (years) | 6.0 (2.0) | 6.4 (1.6) | 13.3 (1.1) | 8.2 (0.7) |

HRT = hormone replacement therapy.
Analyses accounted for sampling design.
